# Dental care of patients with dementia: A survey on practice equipment, training, and dental treatment

**DOI:** 10.1101/2021.03.18.21253826

**Authors:** Sophie Schaper, Sinja Meyer-Rötz, Claudia Bartels, Jens Wiltfang, Tina Rödig, Björn H. Schott, Michael Belz

## Abstract

For patients with dementia, dental care can pose a considerable challenge due to cognitive impairment, behavioral, and psychological symptoms, and (often subsequently) limited autonomous oral care. In this study, we aimed to assess the proficiency of dentists in general practice in the outpatient dental care of these patients. A total of 119 dentists from private practices in Lower Saxony, Germany, participated in this study. Concerning treatment of patients with dementia, they provided details about (1) practice equipment/consultation, (2) training/ expertise, and (3) special circumstances of dental treatment.

Participating dentists predominantly reported to use medical aids (e.g., positioning cushions) to improve the treatment situation for patients with dementia. Over two thirds (68.6%) offered consultations in nursing homes, and at the patients’ homes (47.0%). The training rate was remarkably low in the field of gerodontology for dentists and their practice staff (< 10%), however, 54.5% expressed interest in such training. The majority of dentists reportedly adapted their treatment strategy to the needs of patients with dementia (e.g., communication, inclusion of caregivers, time management). Furthermore, most participants adapted dental treatment adequately (e.g., strict indication for tooth extraction, simple design of dental prostheses).

In summary, even though training in the field of gerodontology must be considered insufficient, most dentists in this study showed an adequate adaptation of their treatment strategy as well as consideration of dental characteristics in patients with dementia, along with interest in trainings. We conclude that dementia-specific training should be expanded in the field of dentistry, preferably already at university level.

## 1 Introduction

Dental treatment can pose a considerable challenge in the growing population of older people (≥ 65 years), as age-related factors like multimorbidity and polypharmacy complicate professional dental treatment (Petersen and Yamamoto, 2005). These circumstances are further aggravated in individuals diagnosed with dementia, the risk of which increases steadily with older age – especially from 65 years and above (Lee et al., 2019; Rosa et al., 2020). The resulting cognitive impairment accompanied by psychological and behavioral symptoms usually prompts the need for long-term dental care. However, patients with dementia are often no longer able to adequately care for their personal health and hygiene, including dental health. Several studies have confirmed a close and possibly reciprocal relationship between dental health and cognitive functioning in older adults (Hatipoglu et al., 2011; Chen et al., 2019; Lauritano et al., 2019). Patients with dementia commonly fail to attend necessary dental appointments and subsequently suffer from a pronounced decline in oral health, including the loss of teeth (Fereshtehnejad et al., 2018; Lopez-Jornet et al., 2021). In addition, poor oral health in patients with dementia is potentially aggravated by pharmacological treatment with neuroleptics or antidepressants, which commonly have adverse side effects like xerostomia or orofacial dyskinesia resulting in bruxism, which in turn increase the need for adequate dental treatment (Fratto and Manzon, 2014).

In addition to lack of dental self-care, patients with dementia also experience increased difficulties with respect to dental interventions. On the one hand, access to dentists may be limited, as most work in private practices, which may be difficult to access for severely cognitively impaired individuals (Fereshtehnejad et al., 2018). Often, the need for dental intervention is not even detected in patients with dementia. For example, when the affected individuals are unable to express themselves adequately and develop agitation symptoms or aggressive behavior due to intraoral pain, the behavioral problems are frequently treated with psychotropic drugs, but the correct detection and/or elimination of the pain’s cause often remains absent (Cohen-Mansfield and Lipson, 2002). On the other hand, dental procedures themselves can prove challenging in older people with dementia. Dementia is often accompanied by pronounced affective and behavioral disturbances, including depressed mood and aggressive behavior, both of which can complicate medical, including dental, treatment (Chalmers, 2000). In more advanced stages of dementia, patients may no longer be cognitively capable of understanding the need for medical or dental visits. In such cases, a relative, health care proxy or legal guardian must be involved, which increases the effort required for treatment and requires the scheduling of longer session times (Niessen and Jones, 1987). Adapting communication to the needs of individuals with reduced cognitive ability (e.g., by speaking in simple, short sentences, keeping eye contact, or communicating via third parties), as well as involving familiar caregivers, can prove challenging in everyday dental practice. At the organizational level, practitioners also face an increased burden, requiring, for example, adequate recall structures to avoid missed appointments and to better monitor oral health. Finally, yet importantly, dental treatment itself may also need to be adapted, owing to both impaired understanding of the required procedures and possible difficulties with day-to-day handling of dentures. One example would be the early switch to a robust, easy-to-care-for and easy-to-handle denture (Nitschke et al., 2021) in order to preserve the functional ability of the masticatory organ as long as possible.

While the problem of dental care in patients with dementia is nowadays widely recognized, less is known about the preparedness among practitioners to adapt their treatments to older people with dementia, and about both, existing approaches and unmet needs in dementia-sensitive dentistry. In the present study, we aimed to assess the status of dental care for patients with dementia by dentists in private practice in southern Lower Saxony, Germany. The region investigated covered a predominantly rural area surrounding the city of Göttingen, encompassing the counties of Göttingen, Goslar, Holzminden, and Northeim (**Figure 1**). The survey reported here is specifically directed at dental care for patients with dementia.

**Fig. 1:**
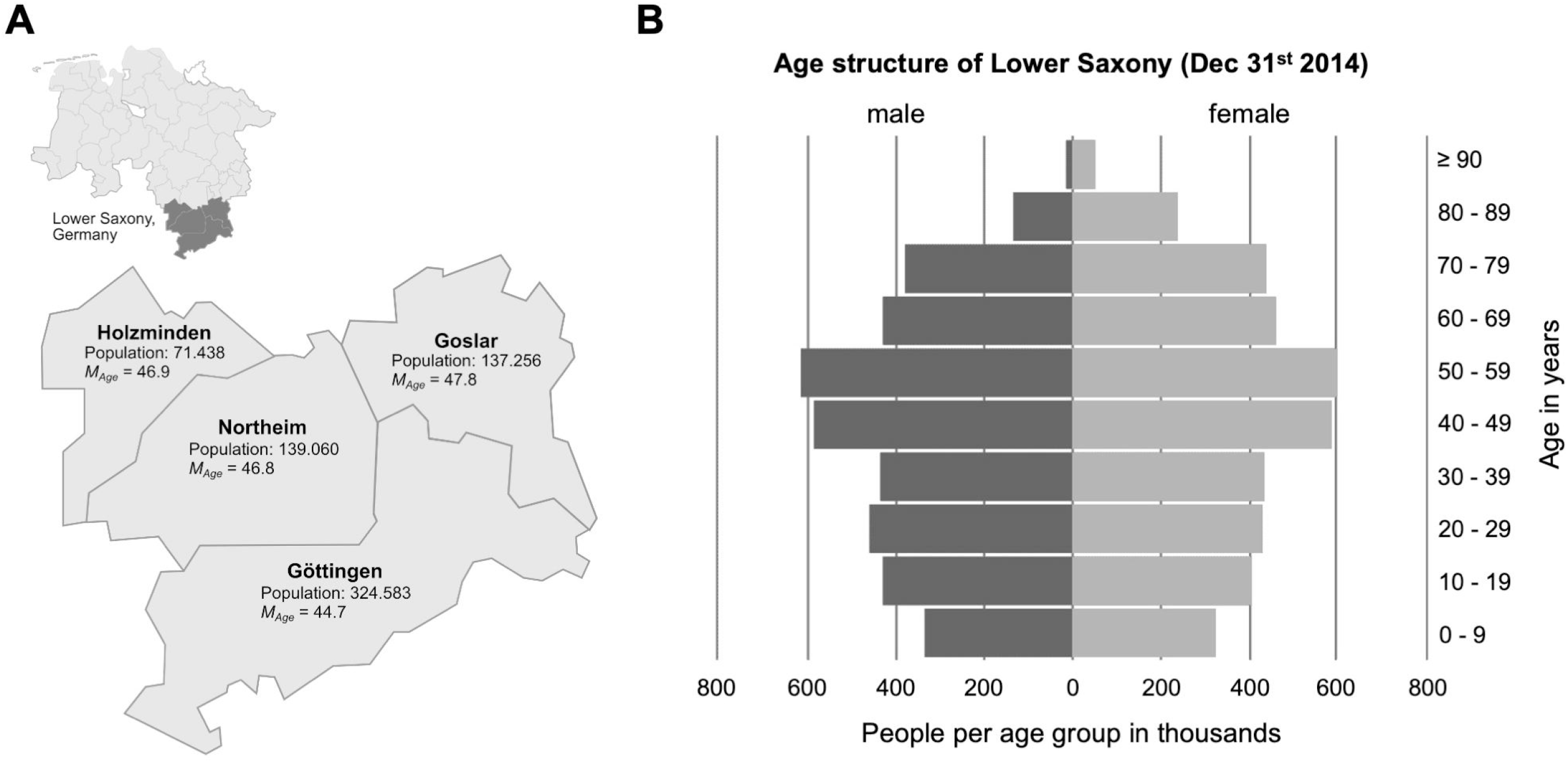
**A:** Display of the region in Lower Saxony, Germany, investigated in this study. The data on the number of inhabitants and mean age of each county are predictions for the year 2014, based on the German population survey “Zensus” from 2011. **B:** Age structure of the State of Lower Saxony in 2014, as provided by the Landesamt für Statistik Niedersachsen (2016). The source materials for generating the map and the age pyramid were obtained from publicly available materials on https://www.niedersachsen.de.

We carried out a mail-based survey, and obtained questionnaire data from 119 dentists in private practice, which were empirically analyzed to address the following three research questions:

1. Are there practice equipment and consultation procedures adequately adapted to treat patients with dementia?
2. Are dentists trained in dementia-specific aspects of gerodontology, and can they rely on existing expertise?
3. What are the special circumstances of dental treatment of patients with dementia, and how are they addressed in practice?

With a response rate of almost 30%, we received a sufficiently large number of completed questionnaires to obtain representative information on dentists’ experience and proficiency in providing dental care to older individuals affected by dementia.

## 2 Methods

### 2.1 Sample and study design

A questionnaire was developed for the present study (see Section 2.2 for details). A total of 406 dentists in private practice enlisted with the Lower Saxony Chamber of Dentists (Zahnärztekammer Niedersachsen, ZKN) were identified in the counties of Göttingen, Holzminden, Goslar, and Northeim, as defined by zip code, and the survey was provided to all dentists by mail in this cross-sectional study. The response rate during the survey period from February 1^st^ 2017 to March 15^th^ 2017 was 29.3% (*N* = 119). Approximately two thirds of the responding participants were male (68.9%, *n* = 82), and the mean age of respondents was *M* = 52.95 years (*SD* = 10.55). The state exam was taken between 1990 and 1991 (*M* = 1990.75, *SD* = 10.42 years), and the mean interval between exam and interview time was 26.25 years (*SD* = 10.42). Female dentists had taken the state exam on average in the year 1996.46 (*SD* = 8.85) compared to male dentists (year: 1988.14, *SD* = 10.07, *t*(116) = 4.32, *p* < .001). They were significantly younger (*M* = 46.89, *SD* = 9.18) compared to their male colleagues (*M* = 55.71, *SD* = 10.01, *t*(113) = 4.49, *p* < .001). The majority of dentists surveyed worked in solo practices (58.8%, *n* = 70), and the remaining respondents worked in group practices (41.2%, *n* = 49). On average, participants reported treating approximately ten individuals with dementia (*M* = 9.51, *SD* = 13.96) per quarter in their practice^1^. All data processing was performed in accordance with European, national, and state regulations on data protection, and the local ethics committee of the University Medical Center Göttingen approved the study.

### 2.2 Measurement

In addition to the collection of basic demographic data, we administered a self-developed questionnaire. It contained a total of 24 items formulated as statements with the dichotomous response options “Yes” and “No” (e.g., “Is your practice barrier-free?”), grouped according to the three research questions. The participating dentists were informed in an accompanying letter that participation in the survey was voluntary and without financial compensation. The survey was part of an empirical study focused specifically on the dental care of patients with dementia. The questionnaires were returned using a stamped envelope provided along with the questionnaire, and data collection was performed in a fully anonymized way. It has therefore at no time been possible to identify any of the participating dentists or their patients. Table 1 shows the allocation of the individual items to each research question: (1) practice equipment and consultation (items 1a to 1h), (2) training and expertise (items 2a to 2i), (3) dementia-related special circumstances of dental treatment (items 3a to 3i). Two open questions were included (item 1e, question (1): use of aids; item 2i, question (2): special features of consultation; see Table 1 for details). Assigned to research question (3), a total of nine ratings on the most frequent intraoral diseases were additionally asked using a 5-point Likert scale (item 3j): “Do you notice intraoral abnormalities and diseases more frequently in patients with dementia?” (e.g., “caries”; response: 1 = “does not apply”, to 5 = “fully applies”).

**Tab. 1:** Questionnaire-items about patients with dementia

| (1): Practice equipment and consultation | Response options |
| --- | --- |
| *1a* "Is your practice barrier-free?" | "Yes" vs. "No" |
| *1b* "Does your practice have handicap accessible restrooms?" | "Yes" vs. "No" |
| *1c* "Do you use assistive devices, such as a pillow to improve positioning?" | "Yes" vs. "No" |
| *1d* "Do you use a dental bench in functionally impaired patients?" | "Yes" vs. "No" |
| *1e* "Do you use any other assistive devices not listed?" | open question |
| *1f* "Consultation in your practice?" | "Yes" vs. "No" |
| *1g* "Consultation in the nursing home?" | "Yes" vs. "No" |
| *1h* "Consultation at patient's home?" | "Yes" vs. "No" |
| (2): Training and expertise | Response options |
| *2a* "Do you have any special training in the field of gerodontology?" | "Yes" vs. "No" |
| *2b* "Does your staff have special training in the field of gerodontology?" | "Yes" vs. "No" |
| *2c* "Do you use any special visual aids, such as flyers/viewing materials, in communication with patients?" | "Yes" vs. "No" |
| *2d* "Does communication with patients occur through third parties (e.g., caregivers, family members)?" | "Yes" vs. "No" |
| *2e* "Can you schedule more time for treatments?" | "Yes" vs. "No" |
| *2f* "Do you set a specific recall structure in advance?" | "Yes" vs. "No" |
| *2g* "Do you use special screening or special tests to detect dementia?" | "Yes" vs. "No" |
| *2h* "Would you be interested in training on how to manage patients with dementia?" | "Yes" vs. "No" |
| *2i* "Are there any special considerations during therapeutic conversation with patients?" | open question |
| (3): Dealing with special circumstances of dental treatment | Response options |
| *3a* "Do you make the indication for tooth extraction stricter in motor-impaired patients?" | "Yes" vs. "No" |
| *3b* "Do you value a prosthetic design that is as functionally simple as possible?" | "Yes" vs. "No" |
| *3c* "Do you switch sooner from fixed to removable dentures?" | "Yes" vs. "No" |
| *3d* "Do you recommend any special oral hygiene products?" | "Yes" vs. "No" |
| *3e* "Do you recommend mouth rinses?" | "Yes" vs. "No" |
| *3f* "Do you recommend a particular brushing technique?" | "Yes" vs. "No" |
| *3g* "Do you treat patients with low compliance with sedating medications?" | "Yes" vs. "No" |
| *3h* "Do you treat patients under general anesthesia?" | "Yes" vs. "No" |
| *3i* "Do you refer patients for inpatient care under general anesthesia?" | "Yes" vs. "No" |
| *3j* "Do you notice intraoral abnormalities and diseases more frequently in patients with dementia?" | Likert-scale<sup>1</sup> |

Notes. <sup>1</sup>Likert-scale (1 = "does not apply" to 5 = "fully applies"). Item 3j could be answered for a total of nine intraoral abnormalities and diseases: (1) caries, (2) gingivitis, (3) pressure sores, (4) periodontitis, (5) denture stomatitis, (6) xerostomia, (7) hypersalivation, (8) mucosal inflammation, (9) carcinogenic changes.

### 2.3 Statistic analysis

Statistical analysis was performed using SPSS version 26 (IBM Corp. Armonk, NY). For descriptive presentation of the questionnaire data, frequencies and percentages were calculated for dichotomous items. Only valid percentages are reported in figures and text, excluding missing values. For dichotomous items, exploratory inferential statistics were performed using multiple chi-square tests (χ^2^). Furthermore, ratings on intraoral disease in patients with dementia are presented as means and standard deviations (*M* ± *SD*), along with the respective 95% confidence intervals (CIs). Differences between these nine intra-individual ratings were tested using a general linear model for multiple measures (GLM). All *p*-values were adjusted for the number of statistical tests within the GLM using the Bonferroni method (initial significance level was set at *p* < .05, two-tailed). Responses to open-ended questions were categorized and then analyzed descriptively (i.e., frequencies).

## 3 Results

### 3.1. Practice equipment and consultation

The valid percentages for the dichotomous items are summarized in **Figure 2A**. While the majority of respondents affirmed the presence of barrier-free access to their practice (63.0%, *n* = 75), handicap accessible restrooms were available in only 31.9% of cases (*n* = 38). The use of assistive devices for positioning (e.g., pillows) was affirmed by a sizable majority (83.9%, *n* = 99), while only 39.1% (*n* = 45) of respondents reported using dental benches for cognitively impaired patients. In response to the open-ended question about other assistive devices (item 1e), *n* = 4 participants responded (“bite wedges,” “patience,” “holding and supporting by staff,” “metal finger guards”). Toilets accessible to the disabled were significantly more common in group practices than in individual practices (χ2(1, *N* = 119) = 4.57, *p* = .045).

**Fig. 2:**
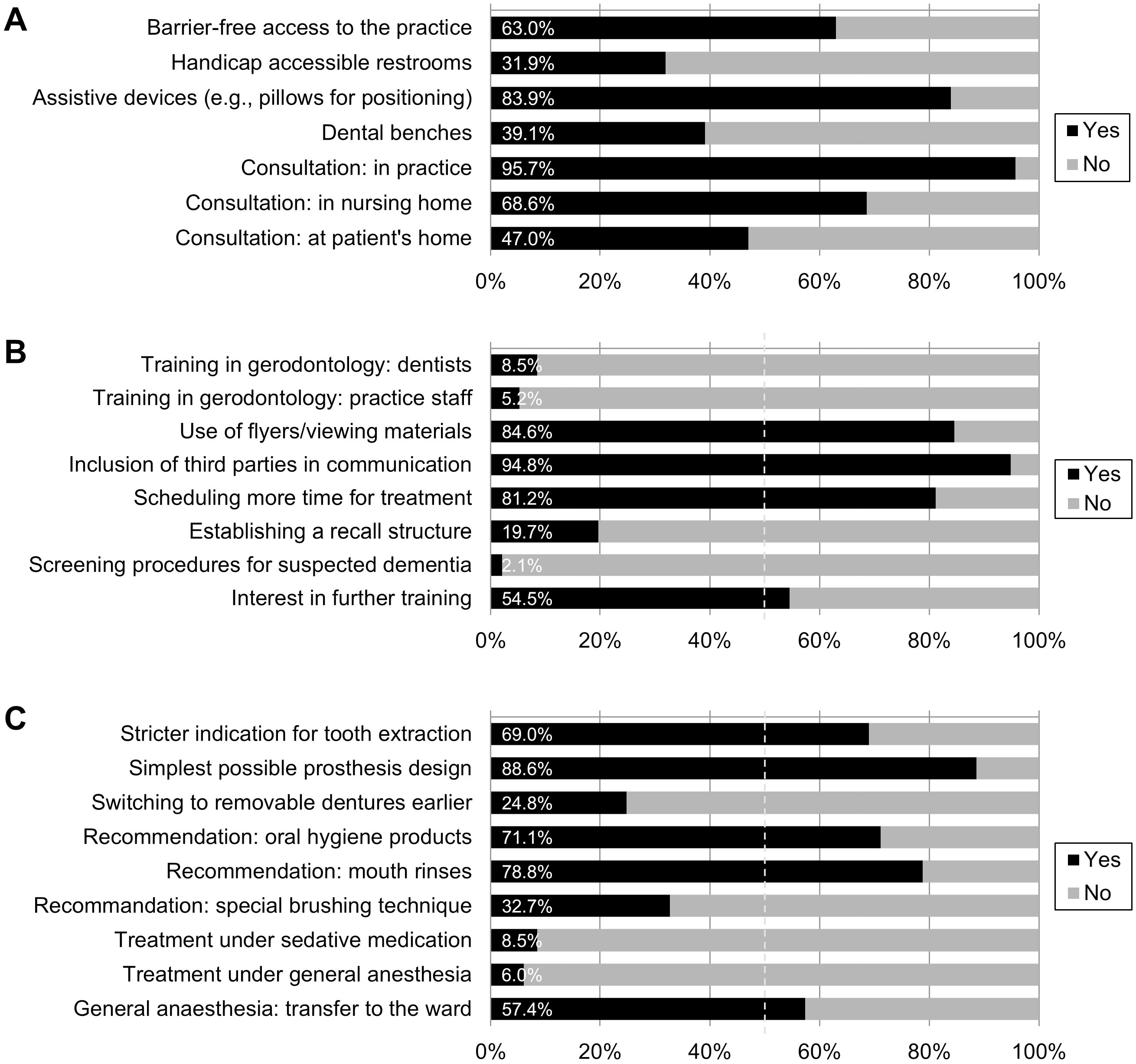
**A:** Practice equipment and consultation. Valid percentages for seven dichotomous items (see Table 1 for formulations), *n* = 100 to 119. **B:** Training and expertise. Valid percentages for eight dichotomous items (see Table 1 for formulations), *n* = 94 to 117. **C:** Dealing with special circumstances of dental treatment. Valid percentages for nine dichotomous items (see Table 1 for formulations), *n* = 104 to 117.

Almost all respondents reported consulting patients with dementia in their practice (95.7%, n = 110), followed by consultation in nursing homes (68.6%, n = 72). Slightly less than half of the dentists reported also seeing patients with dementia at home (47.0%, n = 47). No further differences were found between the two types of practice.

### 3.2 Training and expertise

**Figure 2B** displays responses to questions assessing training and expertise. Only a small minority of the participating dentists (8.5%; *n* = 10) stated that they had received further training in the field of gerodontology, and among the practice staff, this number was even lower (5.2%; *n* = 6). Despite the lack of specific training, the majority of respondents affirmed the use of flyers/viewing materials in communication with patients with dementia (84.6%, *n* = 99), the inclusion of third parties such as relatives in therapeutic conversation (94.8%, *n* = 110), and scheduling more time for treatment (81.2%, *n* = 95). A recall structure in advance of treatment was established by 19.7% of the participating dentists (*n* = 23). Only two respondents (2.1%) used screening procedures for suspected dementia. More than half of the respondents expressed interest in further training in dealing with patients with dementia (54.5%, *n* = 61). Analyses did not reveal significant differences between the types of practice (single practice vs. group practice) for any of these items.

A total of *n* = 26 participants answered the free-text question on special considerations during therapeutic conversation with patients with dementia (item 2i), but in some cases repeated information from the previous items in their own words. Beyond this, the majority of comments referred to the way of speaking with patients (*n* = 11: language/word choice “clear, calm, friendly, loud, slow”). Single participants reported paying attention to keeping the course of treatment as consistent as possible to avoid confusing patients (*n* = 3). A total of *n* = 7 participants made reference to conducting the conversation via third parties, describing ideally only “involving” accompanying persons, and conducting the conversation with the patients themselves as far as possible.

### 3.3 Dealing with special circumstances of dental treatment

See **Figure 2C** for a listing of the valid percentages for the dichotomous items. The majority of participating dentists gave a stricter indication for tooth extraction in patients with dementia and concomitant motor impairment (69.0%, *n* = 80) and paid attention to using the simplest possible denture design (88.6%, *n* = 101). At the same time, only a minority of practitioners switched to removable dentures earlier in the course of treatment (24.8%, *n* = 28). The majority of participating dentists recommended specific oral hygiene products (71.1%, *n* = 81) as well as mouth rinses (78.8%, *n* = 82), and 32.7% (*n* = 37) also recommended a special brushing technique. While practitioners in group practices were significantly more likely to recommend special brushing techniques (χ^2^(1, *N* = 113) = 5.21, *p* = .027), practitioners in individual practices were significantly more likely to pay attention to simple prosthesis design (χ2(1, *N* = 114) = 4.43 *p* = .042). No further differences between the two types of practices were found.

Treatment of patients with low compliance under sedative medication was performed by a small number of respondents (8.5%, *n* = 10), as was treatment under general anesthesia (6.0%, *n* = 7). A majority reported to refer patients with dementia to inpatient dental treatment, if general anesthesia was needed (57.4%, *n* = 62).

Overall, participants estimated the queried intraoral diseases in patients with dementia as being differently frequent (GLM: *F*(8, 776) = 103.40, *p* < .001, partial η2 = 0.69, see **Figure 3** for all assessments). They reported the lowest frequency for carcinogenic changes (*M* = 2.04, *SD* = 0.84) and hypersalivation (*M* = 2.59, *SD* = 0.82): both were significantly lower than all remaining diseases (pairwise comparisons: *p* < .001). The value of 3 = “undecided” was exceeded for all other diseases (*M* from 3.21 to 4.29). The intraoral conditions significantly rated as most common in patients with dementia (value ≥ 4, “somewhat agree”) were caries (*M* = 4.06, *SD* = 0.85), periodontitis (*M* = 4.08, *SD* = 0.83), and gingivitis (*M* = 4.29, *SD* = 0.70). In sum, participating dentists assessed the probability of intraoral diseases to occur more frequently in patients with dementia between 3 = “undecided” and 4 = “more likely true” (overall *M* = 3.44).

**Fig. 3:**
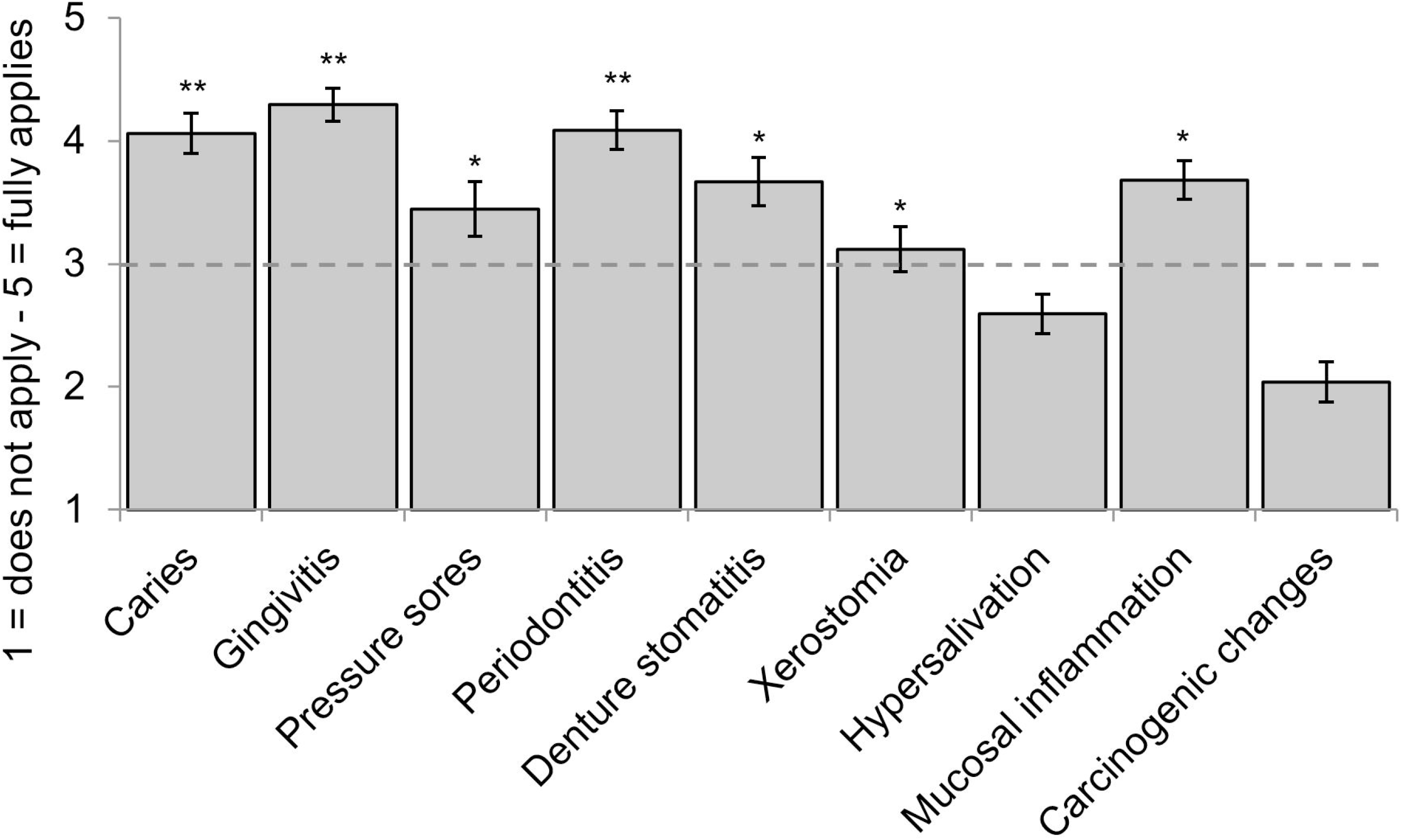
Estimated intraoral diseases in patients with dementia (means, 95%-CIs). Caries, gingivitis, pressure sores, periodontitis, denture stomatitis, xerostomia, hypersalivation, oral mucosal inflammation, carcinogenic changes; *exceeds expression 3 = “undecided”; **exceeds expression 4 = “somewhat agree,” *n* = 101 to 106.

## 4 Discussion

The aim of the present study was to obtain more information on the hitherto insufficiently researched current state of outpatient dental care in patients with dementia. To this end, we evaluated the results of a cross-sectional survey of 119 dentists in private practice, focusing on: (1) practice equipment and consultation, (2) training and expertise, (3) dealing with special circumstances of dental treatment.

### 4.1 Current state of dementia-sensitive dental care

In the majority of practices, aids were used to improve the treatment situation in order to render dental treatment in patients with dementia as stress-free as possible. Deficits in equipment were evident in the use of dental benches and the low availability of toilet facilities suitable for individuals with special needs. Despite the considerable associated time burden, more than two-thirds of the participating dentists reportedly performed consultations in nursing homes, and almost half of the respondents even performed consultations at individual patients’ homes. Although this does not *per se* indicate a high level of continuity of care, it does at least indicate a good quality of care based on studies to date (Knabe and Kram, 1997; Miremadi et al., 2017).

The reported sensitivity for the specific issues related to dental care in patients with dementia was in sharp contrast to the rate of specific training in gerodontology: it was extremely low among both dental and practice staff, with only a small minority of respondents reporting a corresponding qualification at the time of the survey. Topics related to senior dentistry are yet underrepresented in undergraduate study programs (Nitschke et al., 2018), and considering the increasing proportion of older people in the population, improvement is strongly warranted (Stuck and Schimmel, 2021). Encouragingly, a majority of respondents expressed interest in specific training in dementia-sensitive dental care. The respondents also stated that they had already themselves undertaken comprehensive efforts to adapt their treatment strategies to the needs of patients affected by dementia (e.g., adapted communication, inclusion of relatives, scheduling of more time) (Niessen and Jones, 1987). This highlights that dental practitioners are indeed sensitive to the topic and raises the question whether the low rate of specific training may rather result from insufficient availability of adequate training opportunities. While time constraints often limit the implementation of further training at the post-graduate level (i.e., in parallel to the daily practice), it seems promising to offer specific gerodontological courses already at the undergraduate level, which would also sensitize aspiring dentists to this important issue.

Regarding dental treatment itself, the majority of dentists reportedly considered the special circumstances in treating patients with dementia. For example, they stated that indication for tooth extractions was generally stricter, and that dentures were designed as simply as possible, indicating that they paid attention to preservation of functionality and ease of use (Nitschke et al., 2021). Early conversion to removable dentures was performed by only a quarter of the respondents. While we did not explicitly ask about the possible reasons for this, we suggest that they could include increased wearing comfort, better acceptance of fixed dentures by the patients, and protection against loss of removable dentures. Overall, respondents rated the incidence of intraoral disease in patients with dementia as significantly increased. This is in agreement with numerous studies (Gao et al., 2020; Yamaguchi et al., 2021; Zeng et al., 2021). Treatment under sedative medication or even general anesthesia in patients with dementia was performed by only a small proportion of the dentists surveyed. Instead, the majority of patients requiring sedation or general anesthesia were referred to full inpatient treatment, which allows closer monitoring. In our view, this was likely adequate, as the dentists surveyed carefully weighed the associated risks and benefits. As individuals with dementia are particularly prone to suffering from delirium as an acute complication or postoperative cognitive dysfunction (POCD) as a possible long-term consequence of general anesthesia (Fong et al., 2015), any form of deeper sedation must be carefully considered in this vulnerable population, and most dental practices are not equipped to adequately cope with such situations.

### 4.1 Limitations

At 29.3%, the response rate in the present study can be considered good for a postal survey (Sinclair et al., 2012). Nevertheless, we cannot completely exclude potential biases. Most importantly, since the questionnaire had a large number of items and a detailed cover letter, it can be assumed that the majority of respondents who provided information also showed above-average interest in the topic. Thus, some statements (e.g., interest in gerodontological training, adaptation of treatment strategy) are very likely to be positively biased – also due to the mean age of the sample (approximately 53 years).

In our sample, the majority of respondents were male (68.9%), but, on the other hand, this is in line with a nationwide survey for the year 2019 (Lower Saxony: 64.5% male dentists in private practice; Bundeszahnärztekammer, 2019). While we cannot exclude that age and gender distribution of the respondents might have influenced the data reported here, it should be noted that no significant differences between male compared to female dentists were found regarding the main outcomes in this study. Future use of online questionnaires could encourage more, including younger, practitioners to participate, and thus further increase the representativeness of the sample beyond the regional sample reported here. A larger number of participants would also further validate the questionnaire employed. As of now, the cross-sectional study reported here essentially represents a snapshot and allows for a primarily descriptive analysis of the data. A longitudinal version of our survey may, for example, prove useful when applied before and after specific gerodontological training, as it could help to assess the efficacy of such an intervention.

Despite the comprehensive questionnaire used in this study, the inclusion of additional items might, retrospectively, have been helpful to assess to what extend dementia-specific training of dentists can be considered (in)-sufficient. First, dentists’ level of knowledge regarding the relationship between dental health and risk of dementia was not assessed – an important question that should be urgently surveyed in future studies, as the level of knowledge may be a moderator or mediator of other variables assessed here. Additional items may also cover fields in dentistry that require specific training, such as root canal treatment in older patients with multiple systemic conditions (AlRahabi, 2019). Second, questions concerning the usage of specialized practice equipment for safe treatment of older patients (e.g., bite blocks) should be added in a future survey, along with questions assessing the knowledge on a broader spectrum of dental diseases especially in older patients.

A further limitation arises from the fact that the questionnaire study was conducted before the onset of the ongoing SARS-CoV2 pandemic: older people, and particularly those with dementia and/or residing in care homes, are particularly vulnerable to a severe or even fatal disease course of COVID-19 (Panagiotou et al., 2021). Dental care is associated with considerable risk of SARS-CoV2 transmission (Alharbi et al., 2020), and specific precautions are required in vulnerable patients.

Follow-up studies should thus assess how dentists have adapted their treatments of older people and particularly those with dementia as a result of the pandemic.

### 4.2 Conclusion

According to our survey results, dentists already use specific aids and treatment strategies as well as adapted therapeutic conversation when treating patients with dementia. Importantly, they are sensitive to an increased incidence of intraoral diseases in this population and show interest in additional specific training. However, the availability of such training for outpatient dentists is limited. An improvement and expansion of both undergraduate and continuing education opportunities is therefore recommendable, particularly since gerodontology will have a significantly higher relevance due to demographic change. Future studies should systematically assess the effects of gerodontological trainings on outpatient dental care in longitudinal studies, and the questionnaire presented here may constitute a helpful tool in the evaluation of such training interventions.

## Data Availability

The original questionnaire data can be obtained from the authors upon request.

## 6 Author Contributions

Design and conception: J.W., S.Sch., T.R.; Data acquisition and analysis: S.Sch., S.M.-R., M.B.; Formal analysis: M.B.; Writing: S.Sch., C.B., B.H.S., M.B. All authors approved the final version of the manuscript.

## 8 Acknowledgments

We would like to thank all respondents for taking the time and effort to participate in our survey.

## Footnotes

1 No other significant differences were found between the demographic variables recorded. Additional exploratory subgroup analyses, based on the demographic information, were performed for the target variables collected in the questionnaire; however, no significant differences were found between the subgroups.

